# Eating behavior trajectories in the first ten years of life and their relationship with BMI

**DOI:** 10.1101/19003665

**Authors:** Moritz Herle, Bianca De Stavola, Christopher Hübel, Diana L Santos Ferreira, Mohamed Abdulkadir, Zeynep Yilmaz, Ruth Loos, Rachel Bryant-Waugh, Cynthia M. Bulik, Nadia Micali

## Abstract

**Background:** Child eating behaviors are highly heterogeneous and their longitudinal impact on childhood weight is unclear. The objective of this study was to characterize eating behaviors during the first ten years of life and evaluate associations with BMI at age 11 years.

**Method:** Data were parental reports of eating behaviors from 15 months to age 10 years (n=12,048) and standardized body mass index (zBMI) at age 11 years (n=4884) from the Avon Longitudinal Study of Parents and Children. Latent class growth analysis was used to derive latent classes of over-, under-, and fussy eating. Linear regression models for zBMI at 11 years on each set of classes were fitted to assess associations with eating behavior trajectories.

**Results:** We identified four classes of overeating; “low stable” (70%), “low transient” (15%), “late increasing” (11%), and “early increasing” (6%). The “early increasing” class was associated with higher zBMI (boys: β=0.83, 95%CI:0.65, 1.02; girls: β=1.1; 0.92, 1.28) compared to “low stable”. Six classes were found for undereating; “low stable” (25%), “low transient” (37%), “low decreasing” (21%), “high transient” (11%), “high decreasing” (4%), and “high stable” (2%). The latter was associated with lower zBMI (boys: β=-0.79; -1.15, - 0.42; girls: β=-0.76; -1.06, -0.45). Six classes were found for fussy eating; “low stable” (23%), “low transient” (15%), “low increasing” (28%), “high decreasing” (14%), “low increasing” (13%), “high stable” (8%). The “high stable” class was associated with lower zBMI (boys: β =-0.49; -0.68 -0.30; girls: β =-0.35; -0.52, -0.18).

**Conclusions:** Early increasing overeating during childhood is associated with higher zBMI at age 11. High persistent levels of undereating and fussy eating are associated with lower zBMI. Longitudinal trajectories of eating behaviors may help identify children potentially at risk of adverse weight outcomes.

## Introduction

Child eating behaviors have received attention, especially due to their potential association with weight. However, previous cross-sectional and a limited number of longitudinal studies produced inconsistent findings. Previous research has suggested that some eating behaviors are stable across childhood, as indicated by moderate correlations between eating behaviors at age 4 and 10 in English (1) and Dutch samples (2), as well as in younger children, between two and five years (3). However, these studies only had access to two data points, precluding a comprehensive examination stability and change. Some eating behaviors, such as fussy eating, which is the tendency to eat only certain foods and to refuse to try new foods, are common and potentially more transient (1). A previous study reported that one third of children exhibit some fussiness during the first four years of life, but many tend to remit by age six with about 4% being persistently fussy.(5) More recently, a study of the same cohort as discussed in this paper, the Avon Longitudinal Study of Parents and Children, found that mothers indicated that more than half of the children at 15 months were fussy about what foods to eat.(2)

Cross-sectional studies (3–5) have primarily suggested that eating behaviors, such as responsiveness to external food cues or emotional overeating are associated with higher child weight. Other eating behaviors, such as fussy eating and responsiveness to internal satiety cues are associated with lower weight (6–8). However, other cross-sectional studies have not replicated these findings (9, 10). Longitudinally, eating behaviors measured at 5-6 years are weakly associated with body mass index (BMI) at about 6-8 years (11). In earlier ages, between 3 months and 9-15 months, a bidirectional association between child eating and weight has been reported.(12) More recently, the bidirectional association between child eating and later BMI was replicated in a sample of Norwegian children, aged 4 to 8 years(13). Furthermore, children who display fussy eating appear to be at higher risk for developing underweight in childhood, but may be at increased risk for later overweight(14, 15). However, some studies report no or only weak longitudinal relationships (16–18).

Overall, childhood eating behaviors and childhood weight outcomes and the longitudinal development of child eating behaviors remains poorly understood. Longitudinal studies often focus on overall mean scores, ignoring heterogeneity and transience of child eating behaviors. We, therefore, aimed to investigate repeatedly measured eating behaviors in a large population-based birth cohort using latent class modeling to identify longitudinal trajectories during the first ten years of life. Furthermore, we examined their relationship with age- and sex-standardized zBMI at age 11. This age was selected as the outcome measures, due to the proximity to the derived trajectories and to ensure the largest and most representative sample of prepubertal children. Our hypothesis was that persistent EB patterns in childhood would be more strongly associated with child zBMI than transient ones.

## Methods

### Participants

Data from the Avon Longitudinal Study of Parents and Children (ALSPAC), a population based, longitudinal cohort of mothers and their children born in the southwest of England (19, 20) were analyzed. All pregnant women expected to have children between the 1^st^ April 1991 and 31^st^ December 1992 were invited to enroll in the study, providing informed written consent. From all pregnancies (n = 14,676), 14,451 mothers opted to take part; by one year 13,988 children were alive. When the oldest children were approximately 7 years of age, an attempt was made to bolster the initial sample with eligible cases who had failed to join the study originally (referred to as Phase 2), however these participants were not included in these analyses. The phases of enrolment are described in more detail in the cohort profile papers (19, 20). One sibling per set of multiple births, was randomly excluded from these analyses to guarantee independence of participants.

#### Eating behaviors

Repeated measures of parent-reported child eating behaviors were available at a maximum of eight time points around the age of 15, 24, 38, 54, 62, 81, 105, and 116 months. Parents were asked if they were worried about their child overeating (“How worried are you because your child is overeating”), and undereating (“How worried are you because your child is undereating”). The remaining questions probed the child’s tendency to be fussy (“How worried are you because your child is choosy”, “How worried are you because your child has feeding difficulties”, “How worried are you because your child is refusing food”). Parents were given the following response options: “no/did not happen”, “not worried”, “a bit worried” and “greatly worry”. The two top categories (“a bit worried” and “greatly worry”) were combined to avoid very low frequencies. Children who had at least one measure of any of the items were included in the analyses (N=12,048). About half (45%) of the included children had data on all 8 time points and ∼85% had data at least 3 time points.

#### Anthropometric data

Weight and height were measured during clinic visits when the children (N = 4,885) were 11 years old (mean=128.6 months, SD=1.64). Height was measured to the nearest millimeter with the use of a Harpenden Stadiometer (Holtain Ltd.). Weight was measured with a Tanita Body Fat Analyzer (Tanita TBF UK Ltd.) to the nearest 50g. BMI was calculated by dividing weight (in kg) by height squared (in m). Age- and sex-standardized BMI z-scores (zBMI) were calculated according to UK reference data, indicating the degree to which a child is heavier (>0) or lighter (<0) than expected according to his/her age and sex(21). We aimed to relate the trajectories of eating behaviors with zBMI at age 11 years. Children with data on both eating behavior and zBMI were included in the final stage of the analyses (N=4,884). A comparison of the distribution of derived trajectories between participants with and without BMI data at 11 years can be found in eTable 5.

#### Covariates

The following indicators of socioeconomic status of the family were used: Maternal age at birth (years) and maternal education status (A-Levels or higher, lower than A-Levels) and parental occupational status (manual, non-manual labor of the highest earner in the family). Further birthweight (grams) and gestational age at birth (weeks) of the children were also used. The indicators of socioeconomic status were treated as potential confounders for the analyses of zBMI and as predictors of missing data for parent-reported EB data. Details of all data are available through a fully searchable data dictionary at www.bristol.ac.uk/alspac/researchers/our-data.

### Statistical analyses

Analyses were conducted from October 2017 to May 2018 and included two stages in line with the classify-analyze framework (22).

First, Latent Class Growth Analysis (LCGA) was used to identify subgroups (“latent classes”) of children who share the same trajectories of eating behaviors (23). In comparison to Growth Mixture Modelling, an alternative approach to identifying these latent classes, LCGA constrains the variation within each class to zero, reducing the number of parameters and simplifying model estimation (23). LCGA was conducted using Full Information Maximum Likelihood (FIML) (24), incorporating indicators of social class (maternal age, maternal education, and manual or non-manual labor of the highest earner in the family) as auxiliary variables to account for the missingness (including attrition) affecting the longitudinal data, as previously described in ALSPAC (19). FIML assumes data are missing at random (MAR), once these auxiliary variables are accounted for and therefore children with at least one measure of eating behavior at any time point. Analyses were stratified by sex to examine possible effect modification. Stratified results were compared against unstratified using combined data using Likelihood Ratio Tests. As the number of classes is not directly estimated, alternative specifications with increasing number of assumed classes were compared using the following model fit indicators: Akaike Information Criterion (AIC), Bayesian Information Criterion (BIC), adjusted Bayesian information Criterion (adj BIC), selecting the lowest values, and entropy, aiming for the highest. In addition to these model fit indicators, the class size and interpretability of the classes were taken into account as recommended by Muthén (24). After selection of the best number of classes, estimations were repeated using 1000 random starts to avoid local maxima.

In the second stage, participants were allocated to their most likely classes according to their posterior probabilities using the maximum-probability assignment rule (25). These predicted classes were then included as explanatory variables in regression analyses of zBMI scores at age 11, which also controlled for the following a priori confounders: maternal age, gestational age, birthweight, and maternal education at birth. Results are reported in terms of adjusted regression coefficients (β) for each class in comparison to the first (reference) class. Since not all children with eating behavior data had data on zBMI, because of attrition affecting later ages, the characteristics of study participants with/without zBMI were compared in order to assess their representativeness of the original study membership (eTable 5). LCGA was conducted in MPlus Version 8 (26). Regression analyses were conducted in Stata 15 (27). All code is available at https://github.com/MoritzHerle/Patterns-of-child-eating-behaviors-and-later-BMI.

#### Ethical approval

Ethical approval for the study was obtained from the ALSPAC Ethics and Law Committee and the Local Research Ethics Committees. All procedures were performed in accordance with the ethical standards laid down in the 1964 Declaration of Helsinki and its later amendments.

## Results

Summary statistics of the study population at baseline are listed in Table 1. Eating behaviors varied at the different time points (Figure 1). Overeating was uncommon, with the majority of parents reporting that their children never engaged in this behavior (77-85% across the 8 time points). Being fussy about food was the most common child behavior, especially at 54 months, when a fifth of the children were described as fussy to a worrying extent.

**Table 1:** Summary statistics of the baseline characteristics of the study population;

| **Baseline characteristics** |  |  |
| --- | --- | --- |
|  | **N available** | **Mean (SD) or N (%)** |
| **Sex (%boys)** | 12048 | Boys: 6208 (52) |
| **Gestational age at birth (weeks)** | 12048 | 39.45 (1.86) |
| **Birthweight (grams)** | 11902 | 345 (546) |
| **Maternal age (years)** | 12048 | 28.31 (4.86) |
| **Maternal A-Levels or higher** | 11375 | 4158 (37) |
| **Parental non-manual labor**<br>**profession** | 9366 | 7558 (81) |
| **zBMI <sup>a</sup> of children at age 11**<br>**(kg/m<sup>2</sup>)** | 4885 | 0.60 (1.14) |
| **ALSPAC Study** |  |  |

**Figure 1:**
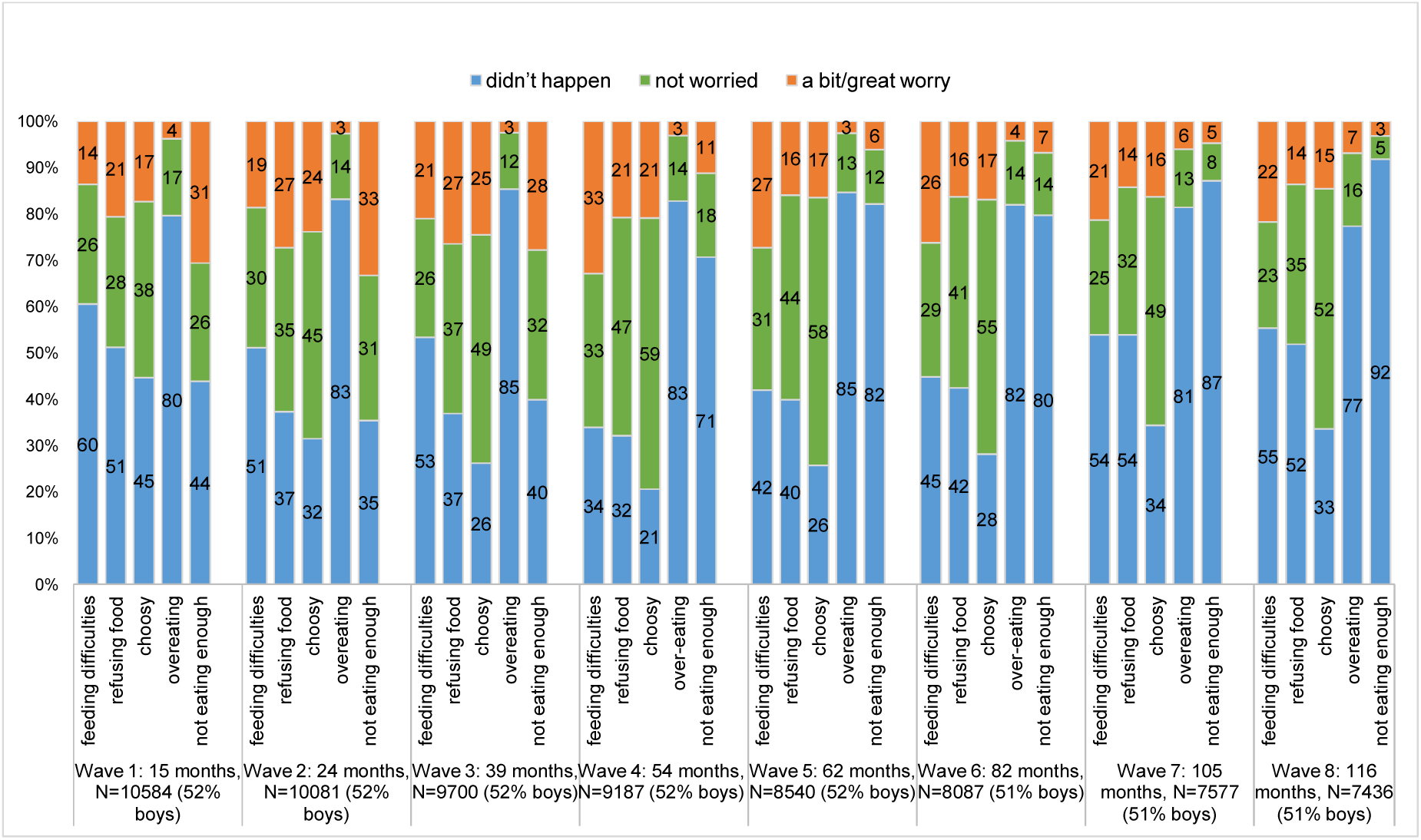
Prevalence of eating behaviours across the eight assessment waves.

### Eating behavior classes

Comparisons of alternative number of classes for the three LGCA models identified four classes for the overeating longitudinal data, and six classes each for undereating and fussy eating according to our pre-specified criteria (eTable 1a-c). Overall, separate models for boys and girls fit the data better than when analyzed jointly (eTable 2).

**Table 2:**
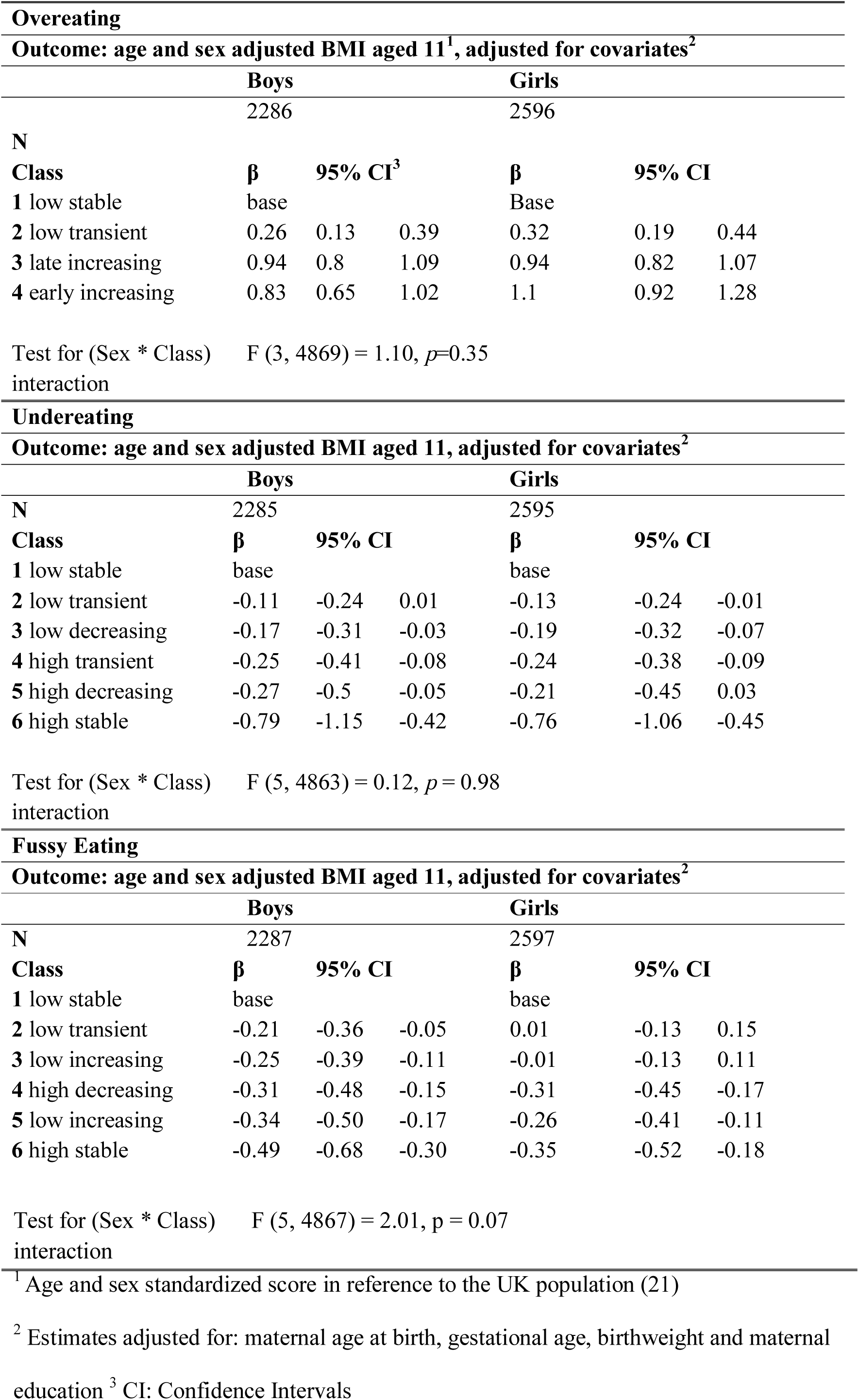
Estimated regression coefficients (β) for assigned eating behavior class on standardized BMI at age 11, separately for boys and girls

Most children were assigned to the “low stable” class of overeating, marked by the absence of high levels of overeating across time points. Undereating was more heterogeneous; the most common class was “low transient”, characterized by low levels of undereating, which attenuated completely by age 10. Similarly, the most common class for fussy eating was the “low transient” group, with increasing numbers of parents reporting fussy eating from 15 months onwards, which decreases again after 62 months. Figure 2a–c illustrates the class trajectories for overeating, undereating, and fussy eating.

**Figure 2a:**
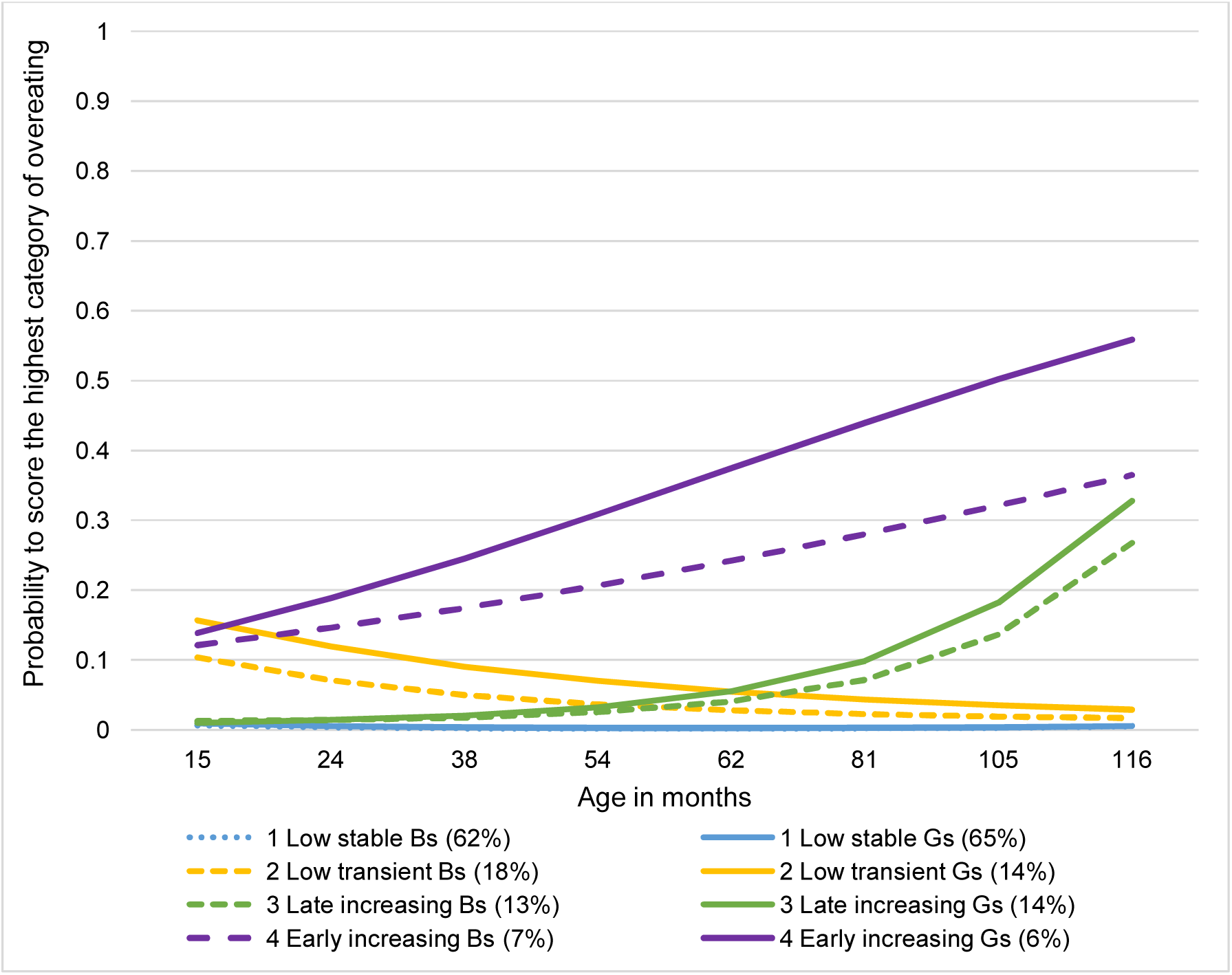
Trajectories of parental reports of overeating behaviors from 15 to 116 months for boys and girls (6186 boys, 5817 girls) **Caption:** The y-axis shows the probability of scoring in the highest category of overeating (“great worry”) at each of the eight time points. Trajectories for boys are in dashed lines. Trajectories for girls are in solid lines. The legend shows the name of the class for boys (Bs) and girls (Gs), followed by their percentages in brackets.

**Figure 2b:**
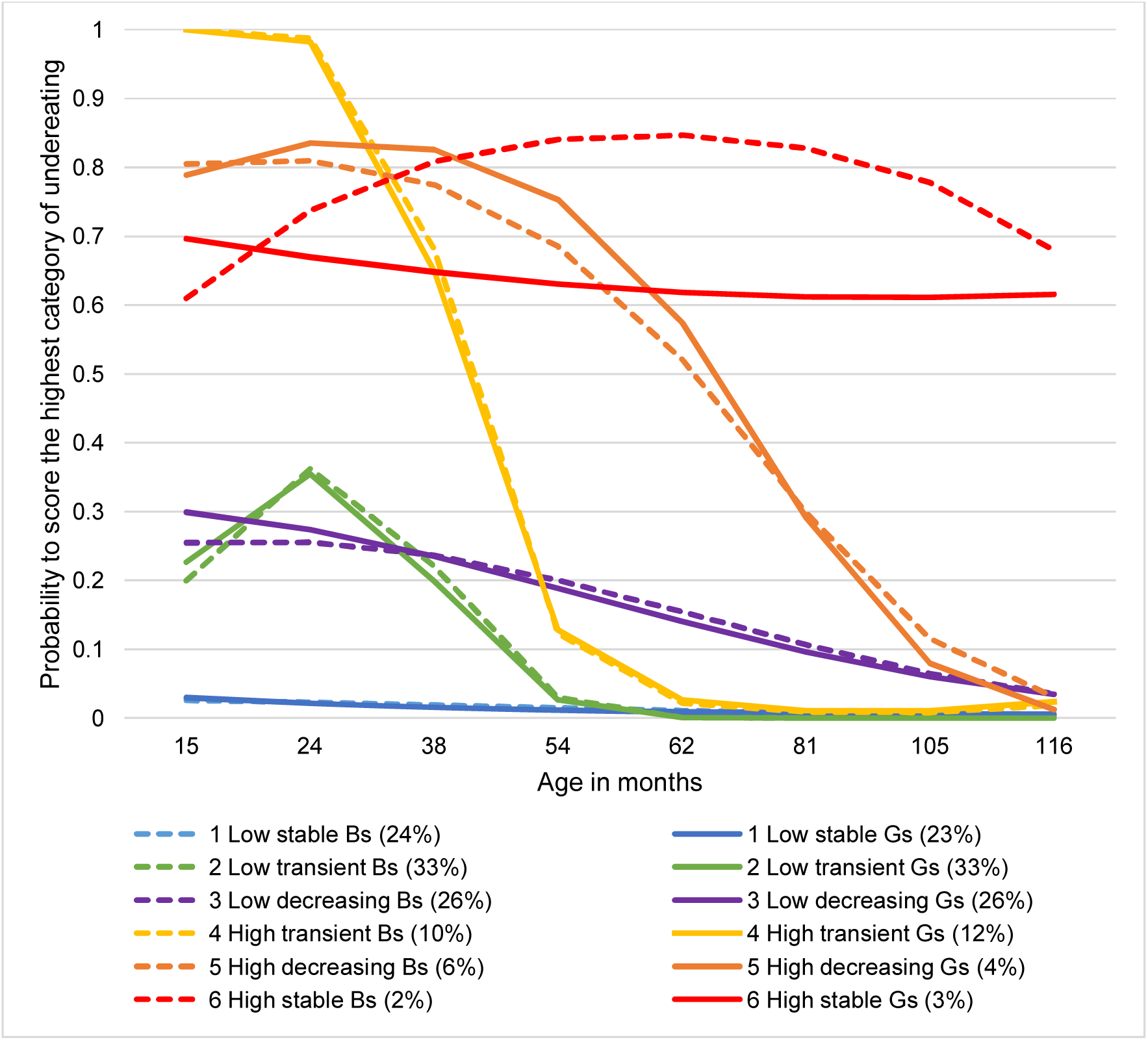
Trajectories of parental reports of undereating behaviors from 15 to 116 months for boys and girls (6189 boys, 5817 girls) **Caption:** The y-axis shows the probability of scoring in the highest category of undereating (“a bit worried”) at each of the eight time points. Trajectories for boys are in dashed lines. Trajectories for girls are in solid lines. The legend shows the name of the class for boys (Bs) and girls (Gs), followed by their percentages in brackets.

**Figure 2c:**
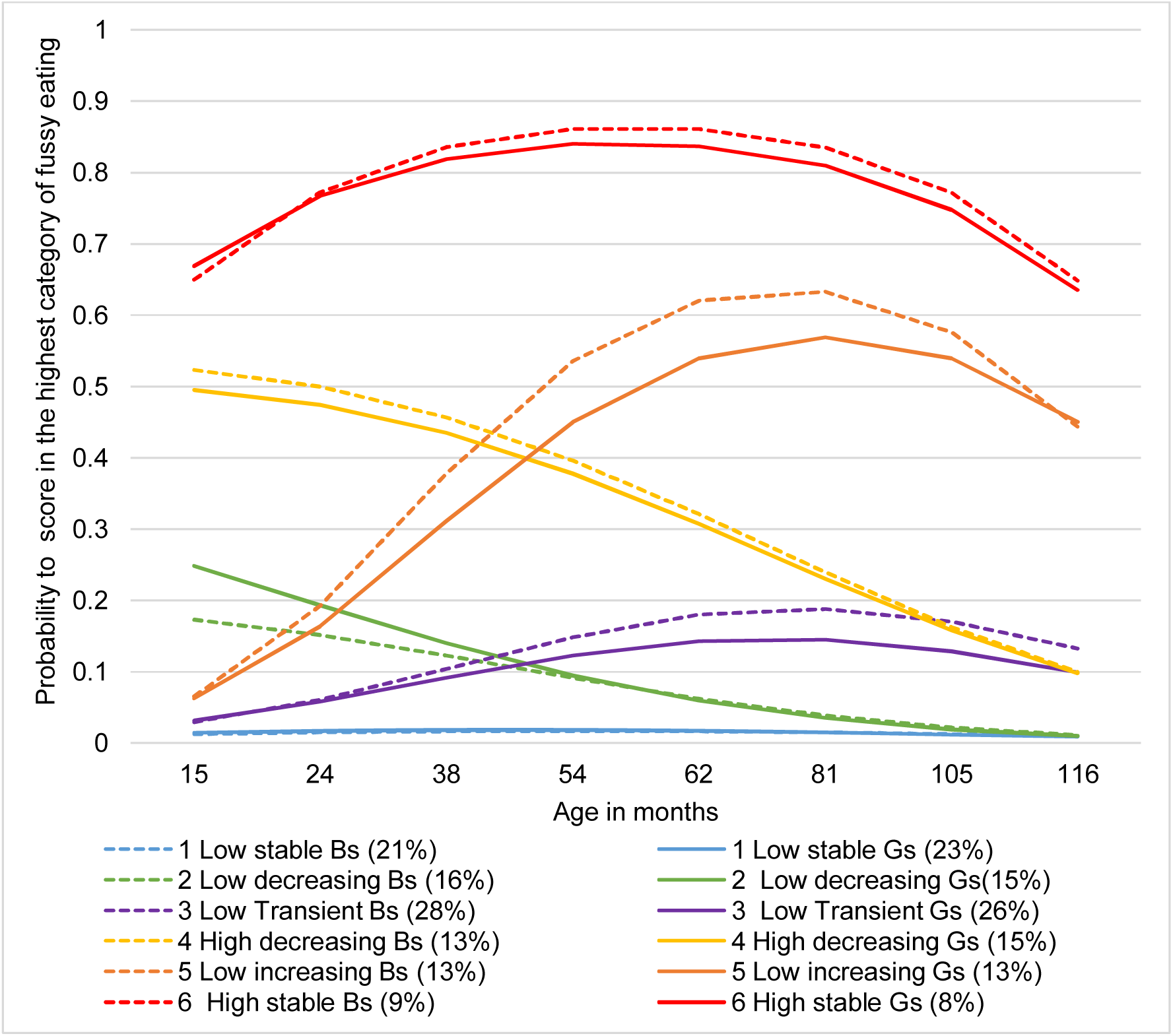
Trajectories of parental reports of fussy eating behaviors from 15 to 116 months for boys and girls (6208 boys, 5840 girls) **Caption:** The y-axis shows the probability of scoring in the highest category of the fussy eating items (“a bit worried””) at each of the eight time points. Trajectories for boys are in dashed lines. Trajectories for girls are in solid lines. The legend shows the name of the class for boys (Bs) and girls (Gs), followed by their percentages in brackets.

When comparing maternal and gestational age at birth, birthweight, and maternal education across classes we found that, for overeating, boys and girls in the “early increasing” class had a higher mean birthweight than those in the “low stable” class; eTable 3). Children in the “high stable” class of undereating and fussy eating had a lower mean birthweight than their respective “low stable” classes. In addition, the percentage of mothers with A-Levels or university degree was lower in the “low stable” class of fussy eating compared to the “high stable”. Child zBMI scores per eating behavior class ranged widely within all classes (eFigure 1a-b).

#### Sensitivity analyses

Not all children included in LCGA had zBMI data at 11 years. Trajectory frequencies derived from all participants were compared to the frequencies among the children who had complete BMI data at 11 years. Trajectory sizes and distributions were similar (eTable 5).

### Association between eating behaviors and zBMI at age 11

#### Overeating

In comparison to children who were reported to never overeat to a worrying extent (“low stable”), all other classes were positively associated with greater zBMI at the later age of 11 years: “low transient” (boys: coefficient β=0.26, 95%CI:0.13, 0.39; girls: β=0.32, 95%CI: 0.19, 0.44), “late increasing” (boys: β = 0.94, 95%CI: 0.8, 1.09; girls: β=0.94, 95%CI: 0.82, 1.07) and “early increasing” (boys: β=0.83, 95%CI: 0.65, 1.02; girls: β=1.1, 95%CI: 0.92, 1.28; Table 2).

#### Undereating

In contrast, undereating classes were associated with lower zBMI. The magnitude of associations in the “high transient” (boys: β=-0.25, 95%CI:-0.41, −0.08; girls: β=-0.24, 95%CI:-0.38, −0.09) and “high decreasing” (boys: β=-0.27, 95%CI:-0.5, −0.05; girls: β=-0.21, 95%CI:-0.45, 0.03) classes were similar. “Stable high” undereating was most strongly associated with lower zBMI (boys: β=-0.79, 95%CI:-1.15, −0.42; girls: β=-0.76, −1.06, −0.45; Table 2).

#### Fussy Eating

Similarly, fussy eating was associated with lower zBMI, for both boys and girls. “Stable high” fussy eating was most strongly and negatively associated with zBMI (boys: β=-0.49, 95%CI: −0.68, −0.30; girls: β=-0.35, −0.52, −0.18; Table 2). In contrast to boys, amongst girls “low transient” and “low increasing” fussy eating were not associated with zBMI at 11 years.

Interactions between class and sex in their effects on zBMI at 11 were not supported for any of the Eating behaviors. Results from unadjusted regression models are available in eTable 6a-c.

## Discussion

In this study, differential developmental patterns in eating behaviors across childhood were identified and found to be associated with later child zBMI. This is the first study to address this question, by establishing longitudinal trajectories of eating behaviors during the first ten years of childhood and investigating their association with childhood zBMI at age 11. Results suggested four different trajectories of overeating and six trajectories each for undereating and fussy eating, respectively. Overall, it is notable that the three eating behaviors follow markedly different developmental trajectories. Overeating was found to be generally low, but increased with time, whereas under und fussy eating varied substantially across the observed timeframe. Previous research, on smaller datasets with a lower number of eating behaviors measures have indicated similar patterns of change and stability (18, 28, 29). These differences might be explained by various complex environmental and biological factors. With age, children gain autonomy and have more meals outside the home, which might be associated with the general increase of overeating. On the other hand, in toddlerhood, parents might introduce various and different textures and flavors of foods to their children (30), which they might readily embrace or resist, potentially explaining this early increase in fussy eating.

The majority of children were not described as overeaters by their parents. However, two trajectories (16% of children) were marked by gradual increases in overeating and showed similar positive associations with child zBMI at age 11. Recent longitudinal analysis of dietary data in a UK child cohort highlighted that eating larger portions a few times per week accelerates early childhood growth (31). This tendency to overeat is likely to result in larger portion sizes, which have been suggested to have enduring effects on child weight (32, 33). Of note, is the possible perpetual bi-directional association between overeating and portion size, where one potentially influences the other.

In contrast to overeating, undereating was more common and more heterogeneous. By 15 months, parents reported that children engaged in various levels of undereating, with about 10% of boys and girls reported to undereat at a worrying level. However, undereating behavior of most children attenuated with time, indicating that parent-perceived undereating in children under the age of two years may represent a normal pattern of development. Only 2-3% of children engaged in persistent high levels of undereating. This persistent pattern of undereating was negatively associated with child zBMI at age 11. Parental reports of undereating might be an indication of satiety sensitivity (34). Previous research has suggested that children who were attuned to their internal satiety cues ate smaller portions (35), and grew at a slower rate than their less satiety-responsive siblings (36).

Similarly to undereating, fussy eating behavior was highly heterogeneous in early life. Using LCGA, we identified a small but substantial group of children (8%) who were persistently fussy throughout the first ten years of life. These results add to previous studies suggesting that some fussiness around food is common during childhood, with one third of children reported to be fussy at some point, but only a small percentage of children remaining highly fussy eaters across development (37). More persistent fussy eating trajectories were negatively associated with child zBMI at age 11.

The relationship between food fussiness and weight is complex as fussy children might undereat certain food groups (e.g., fruits and vegetables) but overeat others (e.g., carbohydrates and fats). Previous cross-sectional studies proposed that fussy children ate fewer vegetables and less fish, but consumed more savory and sweet snack foods at 14 months (38). However, a longitudinal study indicated that persistent fussy eating in childhood was associated with higher prevalence of underweight in children aged six years (15).

This study supports the prospective association between eating behaviors and weight in children. Individual differences in weight have consistently been shown to be influenced by genetic factors (39). The behavioral susceptibility to obesity theory (40) suggests that eating behaviors might act as a mediator between genetic risk for obesity and exposure to the current obesogenic environment. Previous studies proposed that increased genetic risk for obesity is associated with decreased responsiveness to satiety cues, as well as greater responsiveness to external food cues in ten year old twins (41). Subsequent research has replicated these findings in Finnish (42), UK (43) and Canadian adults (44). However, previous studies only included single measures of eating behaviors and it remains unknown how genetic risk for obesity influences longitudinal trajectories of eating behaviors across development.

Apart from weight, eating behaviors have been implicated in diet quality and as potential risk factor for eating disorders. Especially, food fussiness has been associated with poor diet quality,(45) such as low consumption of vegetables (46). Food fussiness has received attention in the context of avoidant/restrictive food intake disorder (ARFID) (47). ARFID is a recently defined diagnosis and little is known about its onset, development, and effect on health and is characterized by extreme food fussiness affecting growth, weight, and physical health (48) and that a large proportion of adolescents diagnosed with ARFID were persistent fussy eaters during childhood (49). More research examining the impact of early food fussiness and undereating on feeding and eating disorders risk is needed. It is possible that the persistent fussy and undereating associated with low zBMI in this study may be ARFID presentations, or risk factors for other eating disorders marked by restrictive eating, such as anorexia nervosa. Further, child food fussiness has been found to be moderately heritable (50) and future research is needed to uncover its genetic basis, as well as the role of fussy eating in neurodevelopmental disorders such as autism spectrum disorder. In addition, the majority of the research in this field relies on parental report. Parental anxiety could influence parents’ perception and reporting of their child’s eating behavior.(51)

## Strengths and Limitations

To our knowledge, this is the most comprehensive longitudinal study of child eating behaviors in a large sample. Data were from a population-based cohort and person-centered statistical analyses allowed us to clarify the heterogeneity of eating behaviors. Height and weight were objectively measured during clinic visits. However, measures of eating behaviors were parent reported and subject to reporting bias. For example parents might be influenced by their own eating behaviors, their prior experiences with other children and might be observing their children more closely in early life. As children grow up and enter school, they will have an increasing numbers of meals outside the family home. Therefore parents might be less aware of their children’s eating behaviors. However, relying on parental report remains the most commonly used measure of child eating behaviors, given that young children are not able to report their own behavior reliably, and standardized direct observational measures are costly and time-consuming, and would be infeasible for large cohorts such as ALSPAC. One additional limitation is the fact that undereating and overeating were only measured with one item at each wave, and a more comprehensive assessment of these eating behaviors would have been desirable. However, in the context of large-scale data collection efforts, such as ALSPAC, researchers always have to strike a balance between including the optimal number of items without overwhelming the participants. Further, the phrasing of the items only enquire how worried parents are about their children’s’ eating behavior, and not the frequency of the behaviors themselves. We implicitly assume that the greater the parental worry, the more pronounced the behavior. The results however refer to the reporting of the behavior, not the behavior per se. Overall, previous support for the use parental reports comes from research validating parent reported child eating against behavioral measures of eating such as eating rate, energy intake at meal, eating without hunger and caloric compensation (35).

Additionally, analyzing the effect of estimated class membership on an outcome includes some degree of uncertainty. The values for entropy, which broadly reflects the level of correct classification, were lower for overeating and undereating than the desired 0.8, commonly used as cut-off point (51). Classes derived from LCGA are unobserved and hence class membership is inferred. We used maximum-probability assignment, which allocates each participant to the class they are most likely to belong to, carrying this class membership forward to further analyses. This method has been suggested to attenuate the effect of class on distal outcomes, due to uncertainty in class assignment (52). Hence, effect sizes estimated from the regression analyses may be conservative.

## Conclusions

We identified four trajectories of overeating and six trajectories each of fussy and undereating in the ALSPAC sample, providing a thorough examination of child Eating behaviors. EB trajectories were differentially associated with child zBMI, with persistent behaviors having a stronger effect on BMI. Characterizing the heterogeneity of early life eating behaviors is an important component to understanding behavioral risk factors for common conditions, such as obesity.

## Data Availability

ALSPAC data access is through a system of managed open access. Please read the ALSPAC access policy (http://www.bristol.ac.uk/media-library/sites/alspac/documents/researchers/data-access/ALSPAC_Access_Policy.pdf) which describes the process of accessing the data and samples in detail, and outlines the costs associated with doing so.

## Acknowledgements

We are extremely grateful to all the families who took part in this study, the midwives for their help in recruiting them, and the whole ALSPAC team, which includes interviewers, computer and laboratory technicians, clerical workers, research scientists, volunteers, managers, receptionists and nurses.

## Author Contributions

MH, BDS, CB, RBW and NM designed the research; MH and BDS performed statistical analyses; all authors wrote and revised the manuscript for important intellectual content; NM had primary responsibility for final content. All authors read and approved the final manuscript.

## Abbreviations

EB: Eating behaviors
BMI: Body Mass Index
ALSPAC: Avon Longitudinal Study of Parents and Children
LCGA: Latent Class Growth Trajectories
ARFID: Avoidant/restrictive food intake disorder

